# Population risk stratification for health systems via accretive predictive modeling

**DOI:** 10.1101/2021.07.19.21260766

**Authors:** Sricharan Bandhakavi, Sunil Karigowda, Zhipeng Liu, Jasmine McCammon, Farbod Rahmanian, Heather Lavoie

## Abstract

**Objective:** Health systems rely on multiple approaches for population-level risk stratification/management. However, they can under-represent members with rising risk and complex treatment needs. To address these gaps and broaden the coverage of members at risk, we present an accretive framework of six predictive models across complementary risk measures for population-level stratification/management.

**Materials and Methods:** Logistic regression models were trained/tested for six outcomes across cost (rising and elevated cost), utilization (rising and elevated utilization), and chronic-disease related (multimorbidities and polypharmacy) risk measures in 2016 using claims-based features from 2015 for ∼8.97 million members in a nation-wide administrative claims database. Model performances were validated against a holdout cohort of ∼2.99 million members. The presence/absence of each outcome prediction for members was summed into an <u>a</u>ccretive <u>p</u>redictive risk index (aPRI) for population-level risk stratification evaluation.

**Results:** Integrating predictions from the six models enabled member stratification across risk measures including future costs, utilizations, and comorbidities. Each of the risk predictions is represented in aPRI levels 0– 6, and their underlying model probabilities/risk measures increase with increasing aPRI levels. ∼83% of members grouped into a “low risk” (aPRI = 0) or “rising risk” category (aPRI = 1 - 2) and ∼17% into a “high risk” (aPRI = 3 - 6) category. Overlap/correlation analyses of risk predictions and comparison of their drivers further support the complementarity of predictions within aPRI and its enhanced coverage of members at risk.

**Discussion:** By integrating targeted and complementary risk predictions, aPRI enhances current population-level risk stratification approaches.

**Conclusion:** We have developed an accretive predictive modeling framework for enhanced population-level risk stratification/management.

## BACKGROUND AND SIGNIFICANCE

Population management broadly refers to the steps taken by healthcare systems and organizations to improve health, cost, and utilization outcomes in a defined group of individuals [1-4]. A fundamental step in this process is the identification of the defined group(s) of individuals via risk stratification approaches and determining the right level of care/services to distinct subgroups of members based on their risk profile. This process is essential to maximize the efficiency of population health programs as a “one-size-fits-all” approach, where the same level of services are offered to every member, is prohibitively expensive and clinically inefficient [5].

Current risk stratification approaches primarily target the identification of members at risk for elevated cost or utilization or chronic disease burden [1-4]. While each of the approaches used has its strengths, they are not without limitations. For example, cost-based stratification approaches can be biased against those with healthcare needs but who are under-utilizers (e.g., racial minorities) [6]. Utilization-based stratification approaches - usually implemented by focusing on members with the highest anticipated use of inpatient or emergency department visits - can under-represent primarily outpatient users who are sizeable contributors to and indicators of overall costs/population health [7 8]. Finally, chronic disease-based stratification approaches are only modestly predictive of future health care utilization/costs [1 9 10], and their ability to capture members with complex treatment needs (e.g. members needing multiple drugs or treatment changes for each disease condition) remains unclear.

An additional limitation associated with all of the above risk stratification approaches stems from focusing on those with the highest risk. Focusing only on high cost high need (HCHN) members can miss members with lower but rising risk who may be more impactable. To target these sub-groups, other groups have developed “impactability” and “propensity to succeed” models for identifying/targeting members more likely to respond favorably with care management or customized interventions [11 12]. However, for a truly population-wide approach to health management, these models must be integrated with the preceding HCHN focused approaches, and this is not always easily achieved.

Thus, for effective population-level risk stratification/management, there is a need to achieve at least two aims: a) address limitations of current approaches aimed mainly at identifying HCHN members and b) allow for easy integration across stratification approaches to retain the value proposition offered by each (approach). Toward these aims, we have developed an accretive strategy based on six complementary predictive models and integrating their predictions for population-level risk stratification/management.

## MATERIALS AND METHODS

### Study design and cohort

We used commercially licensed, nationwide administrative claims data for developing the predictive models described in this study; through the rest of this manuscript, we refer to this data source as the CCAE (<u>C</u>ommercial <u>C</u>laims <u>a</u>nd <u>E</u>ncounters) database [13 14]. Members who were enrolled for both medical and pharmacy benefits through January 01, 2015 –

December 31, 2016, were selected (11.96 million members; aged ≤ 65 years) as the study cohort for this report. The study cohort was processed further by splitting into a train/test dataset (8.97 million members - used for training and testing predictive models described below) and a holdout/validation dataset (2.99 million members - used to validate the performance of the predictive models trained on 8.97 million cohorts, i.e., entire train/test dataset).

### Predictive modeling outcome variables

Six logistic regression-based predictive models were built using scikit-learn/Python [15] to capture risk across cost, utilization, and chronic disease burden. For each predictive model, SQL/python scripts were used to identify members within the corresponding risk/outcome variable of interest in 2016 and labeled as “1” (if the risk was present) or “0” (if the risk was absent). See below for additional details:

a. *Rising cost risk:* members were labeled as having “rising cost risk” if they migrated from a lower (total) cost bucket in 2015 to a higher (total) cost bucket in 2016 or remained in the top ∼1% total cost bucket over 2015-2016. For calculating total costs/cost buckets, paid costs for pharmacy and medical claims were summed for 2015 and cost buckets generated based on ∼80%ile of total costs (< $5000), ∼80-95%ile of total costs ($5000 - $20,000), ∼95-99%ile of total costs (>$20,000 and < $100,000), and >∼99%ile (≥ $100,000). 12% of members had “rising cost risk” in the train/test and holdout datasets.
b. *Elevated cost risk:* members were labeled as having “elevated cost risk” if their total (pharmacy + medical) costs paid in 2016 placed them in the top 10% (> $12,000) of all members. 10% of members had “elevated cost risk” in the train/test and holdout datasets.
c. *Rising utilization risk:* members were labeled as having “rising utilization risk” if they migrated from a lower (total) medical utilization bucket in 2015 to a higher (total) medical utilization bucket in 2016 or remained in the top ∼1% total medical utilization bucket over 2015-2016. For calculating total (medical) utilization claims per member, all their distinct inpatient (IP), outpatient (OP), and emergency department (ED) claims were extracted from medical claims and summed; IP/OP/ED claims were identified using NCQA guidelines for procedure codes [16], place of service codes, and claim type codes. Utilization buckets were generated based on ∼80%ile of total utilization claims (≤ 19 distinct claims), 80-95%ile of total utilization claims (20 - 42 distinct claims), ∼95-99%ile of total costs (43 – 81 distinct claims), and >∼99%ile (> 81 distinct claims). 12% of members had “rising utilization risk” in the train/test and holdout datasets.
d. *Elevated utilization risk:* members were labeled as having “elevated utilization risk” if their total medical utilizations (sum of distinct IP, OP, ED claims identified as described above) in 2016 placed them in the top 10% (i.e., > 31 distinct claims) of all members. 10% of members had “elevated utilization risk” in the train/test and holdout datasets.
e. *Polypharm Rx challenge risk:* Members were labeled as having “polypharm rx challenge risk” if they had ≥3 distinct medication choices or distinct rounds of treatment options for any of five chronic conditions (behavioral health, inflammation/pain, diabetes, hypertension, and chronic respiratory disease) in 2016. Preceding conditions were selected based on their relative prevalence or impact on costs/utilizations. The count of distinct medication choices and rounds of treatment options for each condition was calculated as described previously [13 17]. Members on medications for preceding conditions were identified using their 4^th^ level ATC codes/disease associations as described elsewhere [9 18]. 5% of members had “polypharm rx challenge risk” in the train/test and holdout datasets.
f. *Multimorbidity risk:* Members were labeled as having “multimorbidity risk” if they had ≥2 distinct comorbidities from <u>C</u>harlson <u>C</u>omorbidity Index (CCI) based disease diagnosis groupings [19] in 2016 medical claims. 5% of members had “multimorbidity risk” in the train/test and holdout datasets.

Binary classification logistic regression models were generated to predict which members would have the above risks in 2016 based on features extracted from 2015 claims data for the same members. These features are elaborated further below.

### Predictive modeling features

101 predictors were generated as features using SQL/python scripts identically for test/train dataset vs. holdout dataset using 2015 claims data and used across all 6 predictive models. Features spanned across demographics (age, sex), total costs paid, medical costs paid, pharmacy costs paid, count of distinct IP and ED claims, count of distinct OP claims, count of distinct IP, OP, ED claims, 9 CCI-based diagnosis groups, 31 Rx risk-based medication groups [9], and 6 procedure-code groups corresponding to category 1 CPT codes. Numeric features (e.g., total/medical/pharmacy costs, the sum of IP/OP/ED claims, age) were used both as continuous variables (after log2 transformation to adjust skewness and min-max scaling to maintain all features scaled between 0 to 1) and as categorical features by binning.

### Predictive model characterization and integration

Each predictive model was evaluated via 3-fold cross-validation across performance metrics using train/test dataset (8.97 million members). Subsequently, predictive models were trained on the entire train/test dataset, pickled, and validated against the holdout dataset (2.99 million members) using default probability thresholds. Finally, predictions for members at risk were generated on the holdout dataset using preceding pickled models and adjusted probability thresholds (to increase “rule-in” utility of class 1 predictions) by requiring the satisfaction of the following criteria: minimum specificity > 85%, minimum precision > 25%, and maximum prevalence of class 1 predictions < 3-fold actual prevalence. Each of the member-level model predictions (returned as 0/1 using applied probability thresholds) across all six models were summed as an <u>a</u>ccretive <u>p</u>redictive risk index (aPRI) and characterized for risk stratification/management utility.

## RESULTS

This study was performed in two stages. In the first stage, we generated and characterized six predictive models using nationwide administrative claims data for 2015-2016. All models were built using a common set of 101 features extracted from 2015 claims data and predicted which members had the following risks in 2016: cost risk (rising cost and/or elevated cost risk), utilization risk (rising utilization and/or elevated utilization risk), and chronic disease-related risk (multimorbidity and/or polypharmacy risk). In choosing to develop these six models, we sought to identify high cost high need (HCHN) members across disparate risk measures, capture those missed by an exclusively HCHN strategy, and integrate across predictions to enhance breadth-of-coverage of members at risk. In the second stage, we integrated all six-model predictions into a composite risk measure - referred to as the <u>a</u>ccretive <u>p</u>redictive risk index (aPRI) – and characterized it further for risk stratification/management utility. Results from each of these stages and their underlying analyses are described further in this section.

### Cost risk predictive models

Cost risk predictive models (rising cost risk and elevated cost risk; see materials and methods for definitions) were generated/evaluated using cross-validation on train/test dataset (8.97 million members). Subsequently, predictive models were trained on the preceding cohort of 8.97 million members, pickled, and “cost-risk” predictions generated on holdout dataset (2.99 million members) using optimized probability thresholds; see materials and methods for details. **Figure 1A** shows model performances in train/test dataset (using default probability thresholds) and holdout dataset (using optimized probability thresholds). Using optimized probability thresholds resulted in increased precision and specificity of both models at the expense of model sensitivity/recall. This yielded a balanced accuracy of 63% for rising cost risk predictions and 74% for elevated cost risk predictions on the holdout dataset.

**Figure 1:**
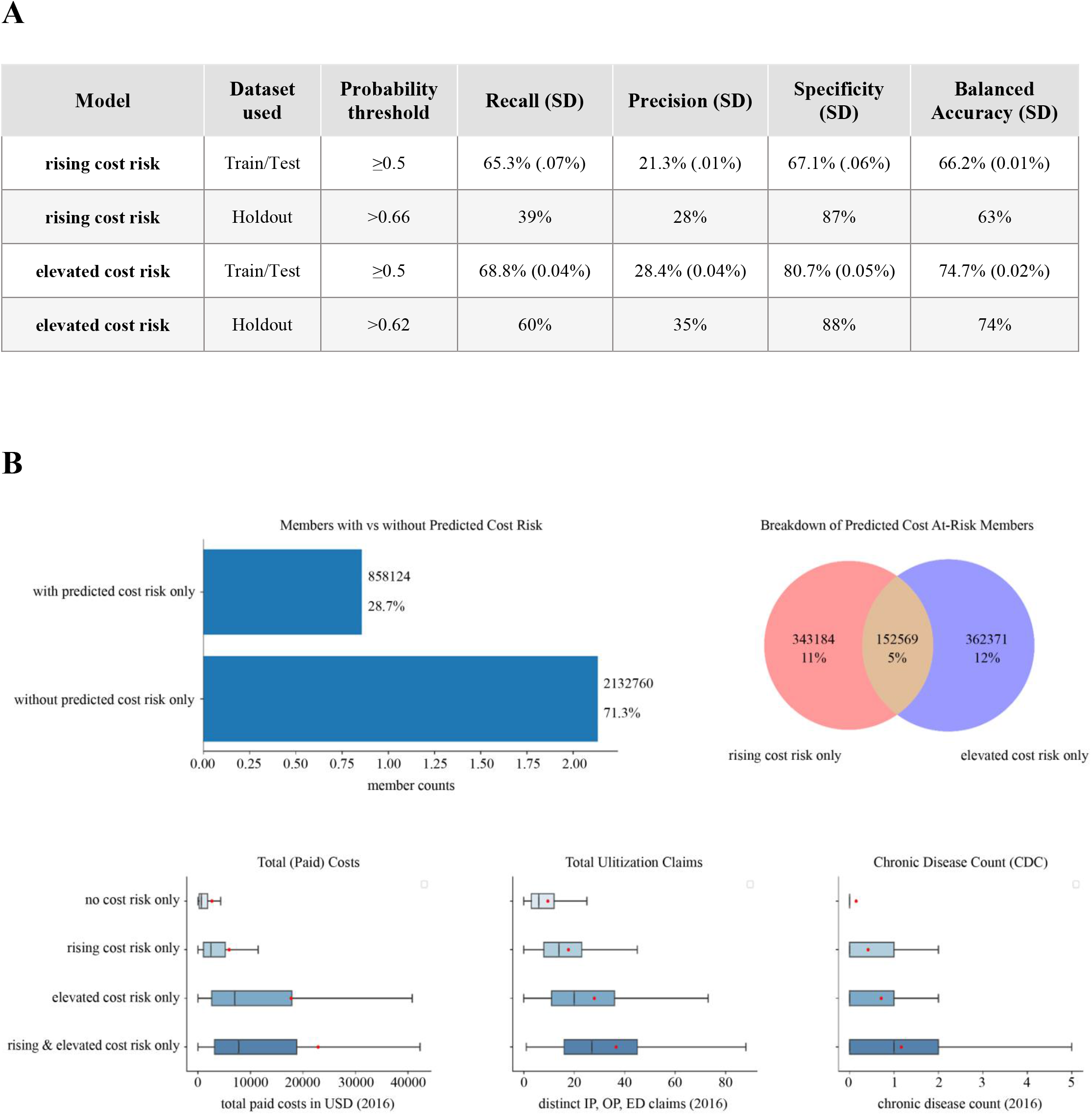
Performance of cost-risk models and characterization of model predictions. **A**. Model performance metrics for next year rising cost risk and elevated cost risk predictions on train/test data (using default probability thresholds) and on holdout data (using indicated probability thresholds). Model performance on train/test data was evaluated using 3-fold cross-validation and mean values across 3-folds are displayed first followed by their standard deviation (SD) values in parentheses for each performance metric. All predictions were based on 2015 claims data for the next year (2016). **B**. Characterization of cost-risk predictions on holdout data for prevalence (top left panel) overlap between future rising cost risk vs. elevated cost risk model predictions (top right panel) and notched box plot distributions of costs, utilizations, and Charlson comorbidity index-based [19] chronic disease counts for each predicted cost-risk group (bottom panel). Notch displays 95% confidence interval around the median, red dot indicates average (mean), and outliers are not shown.

Next, we further characterized each of the cost risk predictions for overlap and utility in risk stratification. As shown in **Figure 1B (top left panel)**, ∼71% of holdout dataset members were not predicted to have either cost risk, and∼29% were predicted to have either rising cost risk or elevated cost risk. Within these ∼29% of members, 11% were predicted to have only rising cost risk, 12% predicted to have only elevated cost risk, and 5% predicted to have both risks (**Figure 1B, top right panel**). Thus, the two cost risk predictive models identify mostly unique members with potentially distinct risk profiles. To evaluate the preceding notion further, we characterized each of the predicted cost risk groups for their 2016 distributions of total paid costs, total utilizations, and cumulative count of selected comorbidities (i.e., are included in Charlson Comorbidity Index). Notched box plot analyses (**Figure 1B, bottom panel)** indicate that across each of these measures, the overall risk increased as follows: “no cost risk only” < “rising cost risk only” < “elevated cost risk only” < “rising & elevated cost risk”, as evidenced by their non-overlapping notched medians for costs, utilizations, and/or comorbidities. Further analysis revealed that among these groups, “rising cost risk only” members had the highest year-over-year averaged cost changes (∼$3151 average *increase*), and “elevated cost risk only” groups had the lowest year-over-year cost changes (∼$5881 average *decrease*) followed by “no cost risk” members (∼$1268 average *increase*) and “rising & elevated cost risk” members (∼$209 average *increase*). Taken together, these results support both the unique nature of each of the cost risk predictions and their utility in risk stratifying members when combined.

### Utilization risk predictive models

Utilization risk predictive models (rising Utilization risk and elevated utilization risk; see materials and methods for definitions) were generated using train/test data, predictions generated against holdout data identically as done for cost risk predictive models, and characterized further detailed in this section. **Figure 2A** shows model performances in train/test dataset (using default probability thresholds) and holdout dataset (using optimized probability thresholds). Using optimized probability thresholds resulted in increased precision and specificity of both models at the expense of model sensitivity/recall. This yielded a balanced accuracy of 61% for rising utilization risk predictions and 78% for elevated utilization risk predictions on the holdout dataset.

**Figure 2:**
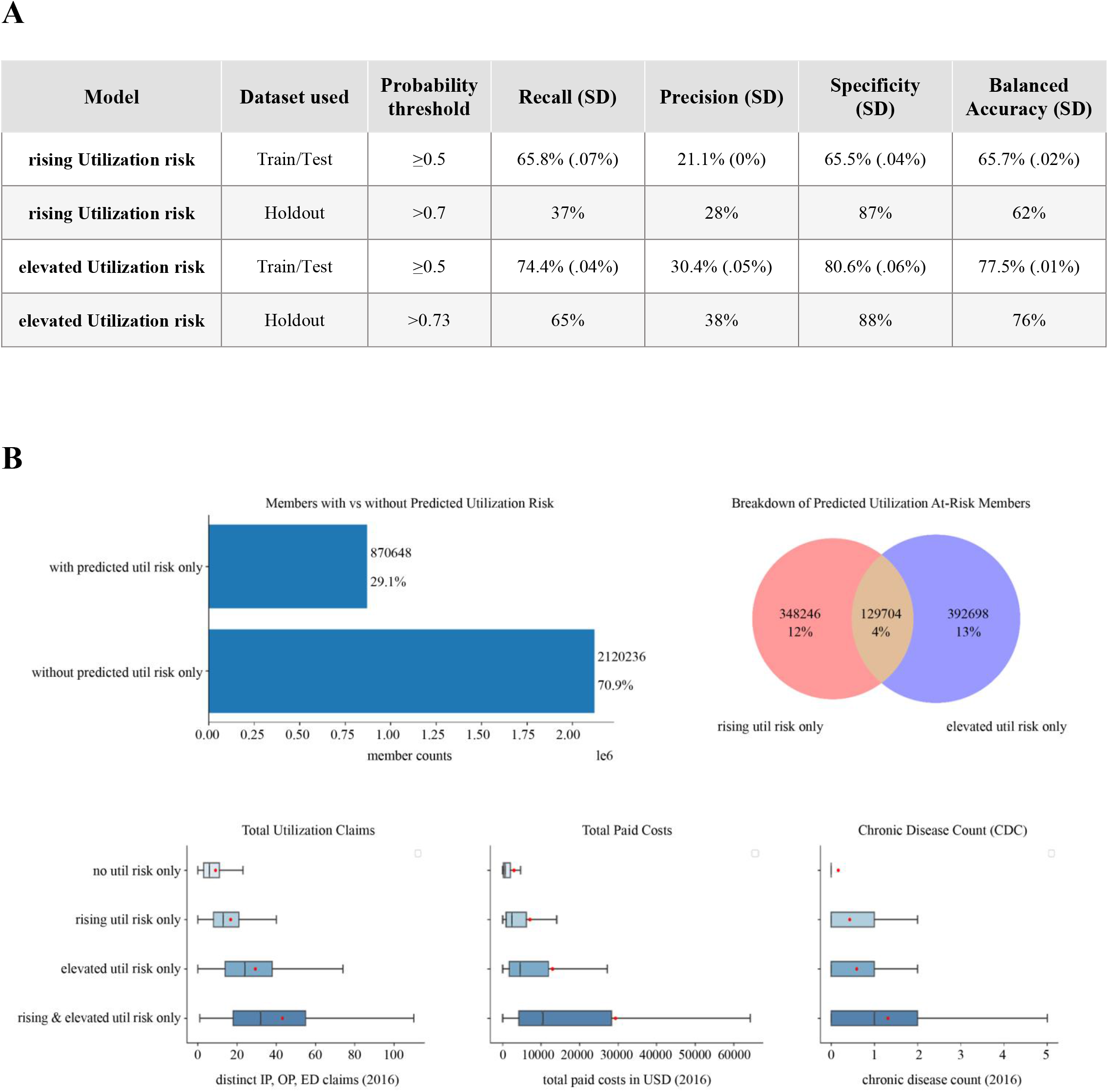
Performance of utilization-risk models and characterization of model predictions. **A**. Model performance metrics for next year rising utilization risk (“rising utilization risk”) and elevated utilization risk (“elevated utilization risk”) predictions on train/test data (using default probability thresholds) and on holdout data (using upwardly adjusted, indicated probability thresholds). Model performance on train/test data was evaluated using 3-fold cross-validation and mean values across 3-folds are displayed first followed by their standard deviation (SD) values in parentheses for each performance metric. All predictions were based on 2015 claims data for the next year (2016). **B**. Characterization of utilization-risk predictions on holdout data for prevalence (top left panel) overlap between future “rising utilization risk” vs. “elevated utilization risk” model predictions (top right panel) and notched box plot distributions of utilizations, costs, and Charlson comorbidity index-based [19] chronic disease counts for each predicted utilization-risk group (bottom panel). Notch displays 95% confidence interval around the median, red dot indicates average (mean), and outliers are not shown.

Next, we characterized further each of the utilization risk predictions for overlap and utility in risk stratification. As shown in **Figure 2B (top left panel)**, ∼72% of holdout dataset members were not predicted to have either Utilization risk andv∼28% were predicted to have either rising utilization risk or elevated utilization risk. Within these ∼28% of members, 13% were predicted to have only rising utilization risk, 10% predicted to have only elevated utilization risk, and 5% predicted to have both risks (**Figure 2B, top right panel**). Thus, the two utilization risk predictions identify mostly unique members with potentially distinct risk profiles. To evaluate the preceding notion further, we characterized each of the predicted utilization risk groups for their 2016 distributions of total utilization, total paid costs, and (selected) total comorbidities. Notched box plot analyses (**Figure 2B, bottom panel)** indicate that across each of these measures, the overall risk increased as follows: “no utilization risk only” < “rising utilization risk only” < “elevated utilization risk only” < “rising & elevated utilization risk”. Analysis of year-over-year utilization claims change revealed the following: “rising utilization risk only” and “no utilization risk only” members had modest year-over-year increases (∼2.2 and ∼2.1 claims on average, respectively) in utilization claims, whereas “elevated utilization risk only” (∼7.9 claims average decrease) and “rising & elevated utilization risk” members (∼4.9 claims average decrease) had year-over-year decreases in utilization claims. Taken together, these results support both the unique nature of each of the utilization risk predictions and their utility in risk stratifying members when combined.

### Chronic disease related risk predictive models

To capture risk associated with chronic disease burden, we generated two predictive models: Polypharm rx challenge (members with at least three distinct Rx choices or rounds of treatment options for any of 5 different chronic conditions, and multimorbidity (members with at least 2 distinct chronic conditions from Charlson Comorbidity Index) risk using train/test data. Subsequently, we generated predictions for members with these risks using holdout data (as done previously for cost risk/utilization risk models). **Figure 3A** shows model performances in train/test dataset (using default probability thresholds) and holdout dataset (using optimized probability thresholds). Optimized probability thresholds resulted in increased precision and specificity of both models at the expense of model sensitivity/recall. This yielded a balanced accuracy of 84% for the polypharm rx challenge and 86% for multimorbidity risk predictions on the holdout dataset.

**Figure 3:**
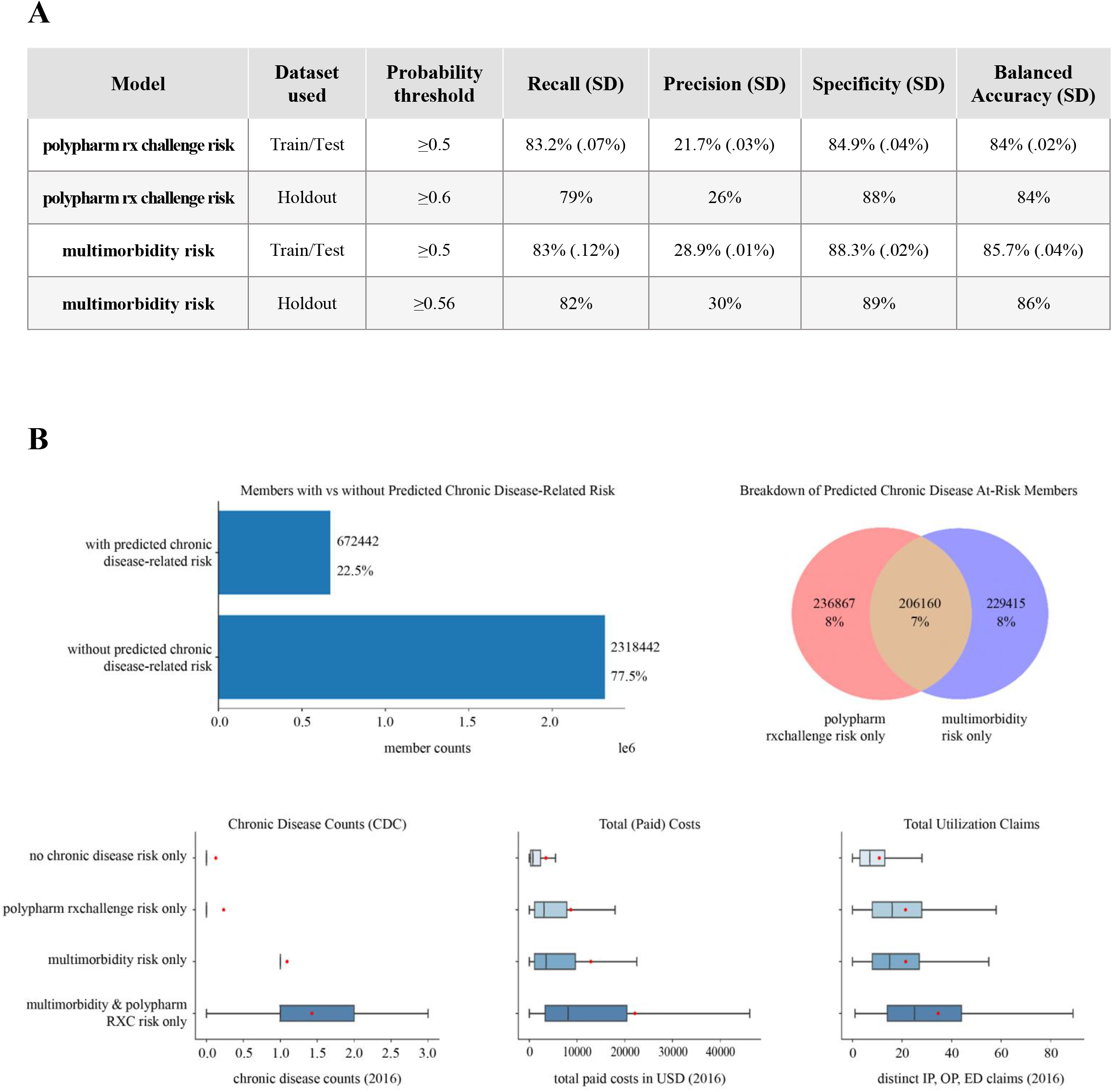
Performance of chronic disease related-risk models and characterization of model predictions. **A**. Model performance metrics for next year multimorbidity risk and polypharmacy rx challenge risk (aka “polypharm RXC risk”) predictions on train/test data (using default probability thresholds) and on holdout data (using upwardly adjusted, indicated probability thresholds). Model performance on train/test data was evaluated using 3-fold cross-validation and mean values across 3-folds are displayed first followed by their standard deviation (SD) values in parentheses for each performance metric. All predictions were based on 2015 claims data for the next year (2016). **B**. Characterization of chronic disease related-risk predictions on holdout data for prevalence (top left panel), overlap between future “multimorbidity risk” vs. “polypharm RXC risk” model predictions (top right panel), and notched box plot distributions of Charlson comorbidity index-based [19] chronic disease counts, costs, and utilization for each predicted chronic disease related-risk group (bottom panel). Notch displays 95% confidence interval around the median, red dot indicates average (mean), and outliers are not shown.

We characterized further each of the chronic disease related risk predictions for overlap and utility in risk stratification. As shown in **Figure 3B (top left panel)**, ∼77% of holdout dataset members were not predicted to have either of the risks, and∼23% were predicted to have either polypharm rx challenge or multimorbidity risk. Within these ∼23% of members, 8% were predicted to have either polypharm rx challenge risk or multimorbidity risk, and 7% predicted to have both risks (**Figure 3B, top right panel**). Thus, the two “chronic disease related risk” predictions identify mostly unique members with potentially distinct risk profiles. Indeed, notched box plot analyses show that (Figure 3B, bottom panel) each of the predicted “at-risk” groups have distinct characteristics across chronic disease counts, total costs, and/or utilization patterns.

### Integration/characterization of combined model outputs for population risk stratification

Following the development/characterization of the six predictive models described above, we proceeded to integrate their predictions into a composite risk score for population-level analyses on our holdout dataset. Towards this goal, we initially evaluated relative prevalence for each prediction and overlap/correlations between risk predictions. Subsequently, each member-level prediction (returned as 0/1 using optimized probabilities as described in methods section) across all six models was summed as an <u>a</u>ccretive <u>p</u>redictive risk index (aPRI). Finally, we characterized the utility of aPRI for population-level risk stratification/management. These analyses are detailed further in this section of the manuscript.

**Figure 4A (top left panel)** shows ∼equivalent representation for each of the six model predictions in the holdout dataset. Prior analysis showed that for each risk measure (cost, utilization, chronic disease), we increased the identification of members at risk by combining two predictive models (**Figure 1B, 2B, 3B, top right panel**). Venn diagrams showed that cost-risk, utilization-risk, and chronic disease-related risk predictions are additionally complementary to each other as well (**Figure 4A, top right panel**), thus enhancing the overall identification of members at risk across these measures when combined. Correlation analyses also support the mutually complementary nature of each of the model predictions (**Figure 4A, bottom left panel**). After summing each of the member-level predictions into an <u>a</u>ccretive <u>p</u>redictive risk index (aPRI), the vast majority of holdout dataset members (>80%) had an aPRI level of 0 - 2, while less than 20% of members had an aPRI level of 3 or higher (**Figure 4A, bottom right panel**). As indicated by notched box plot analyses in **Figure 4B**, increasing aPRI levels are associated with increasing future costs, utilizations, and/or comorbidities. Thus, by integrating each of our six risk predictions into the aPRI, it not only captures a broader set of members at risk but also supports their stratification across multiple risk measures.

**Figure 4:**
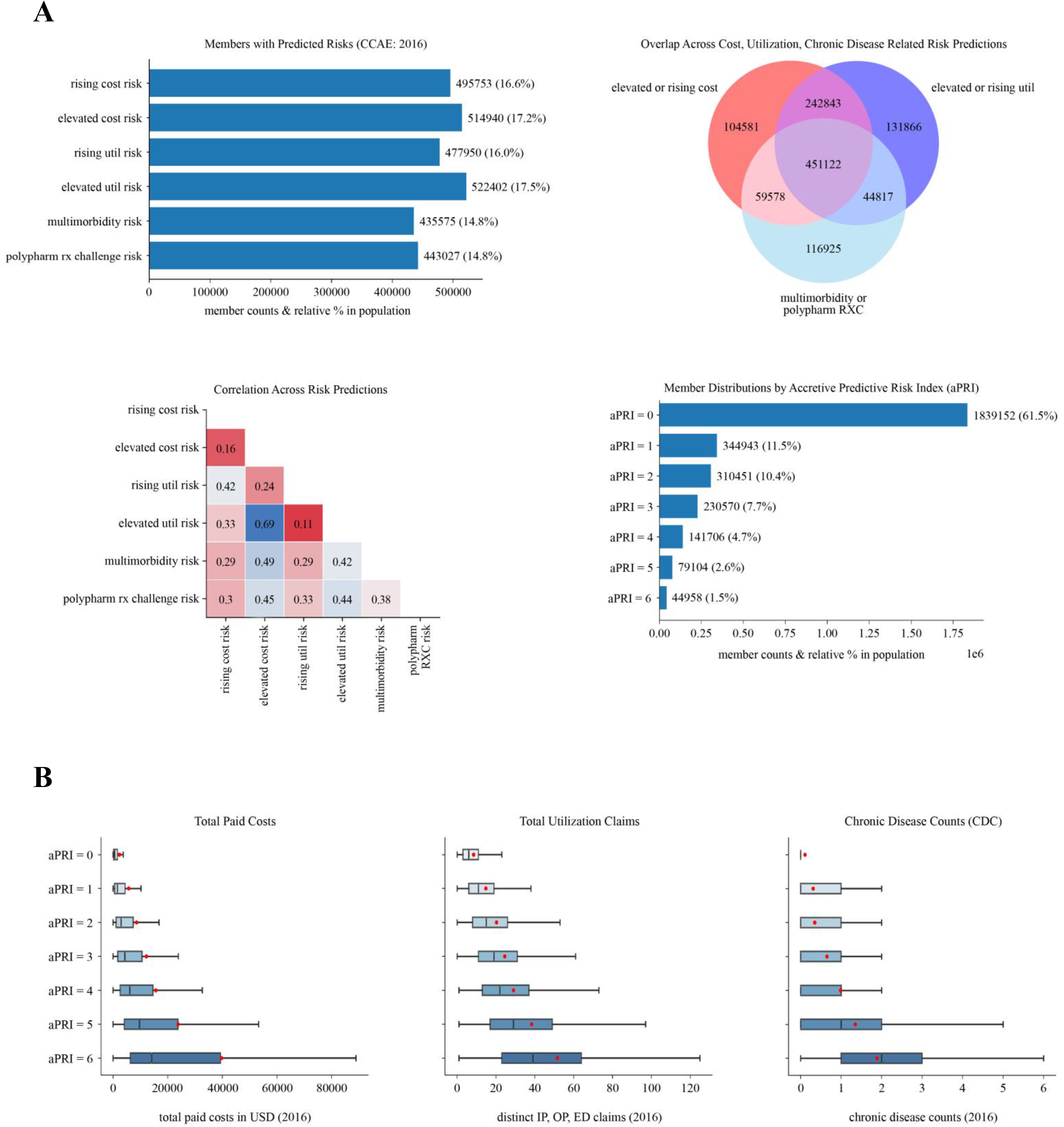
Integration of model predictions via <u>a</u>ccretive <u>p</u>redictive risk index (aPRI) and evaluation of aPRI for risk stratification of members. **A**. Count, relative prevalence of “at-risk” (class 1) predictions from each input model used for calculation of member-level aPRI value (top left panel), Overlay/correlation analysis of at-risk predictions (top right and bottom left panels), distribution of member count, relative percentages for aPRI levels from 0 – 6 (bottom right panel). All characterization was done on holdout data members. **B**. Notched box plot distributions of costs, utilizations, and Charlson comorbidity index-based [19] chronic disease counts for holdout data members with aPRI levels 0 – 6. Notch displays 95% confidence interval around the median, red dot indicates average (mean), and outliers are not shown.

Subsequently, we sought to understand better the ability to stratify across risk measures with increasing aPRI levels for members in the holdout dataset. Towards this aim, we initially evaluated the relative prevalence of each input model’s risk predictions at aPRI levels 1-6 (**Figure 5A**) and later characterized the distribution of input model probabilities by each aPRI level (**Figure 5B**). As shown in **Figure 5A**, each input model’s “at-risk” predictions are represented in aPRI levels 1-6, but there are unique differences in their relative prevalence at each aPRI level. aPRI levels 1-2 have an apparently higher representation of rising cost risk and rising utilization risk predictions (members with lower levels of cost, utilization, and chronic disease counts per **Figures 1B, 2B**) compared to elevated cost/utilization risk and chronic disease-related risk predictions. The latter increase in prevalence in aPRI levels 3-5 and by aPRI level 6, there is ∼equivalent representation of each input model as expected. Notched box plot analyses demonstrate that increasing aPRI levels are also accompanied generally (except perhaps for rising cost risk and rising utilization risk predictions) with increasing probabilities of their input models. Consequently, input model predictions have higher precision (due to higher model probabilities) at higher aPRI levels, further supporting their observed increased costs, utilization, and chronic disease counts (**Figure 5B**). Thus, we propose that relative representation of each input model and the distributions of their model probabilities likely explain the ability to stratify across risk measures with increasing aPRI levels.

**Figure 5:**
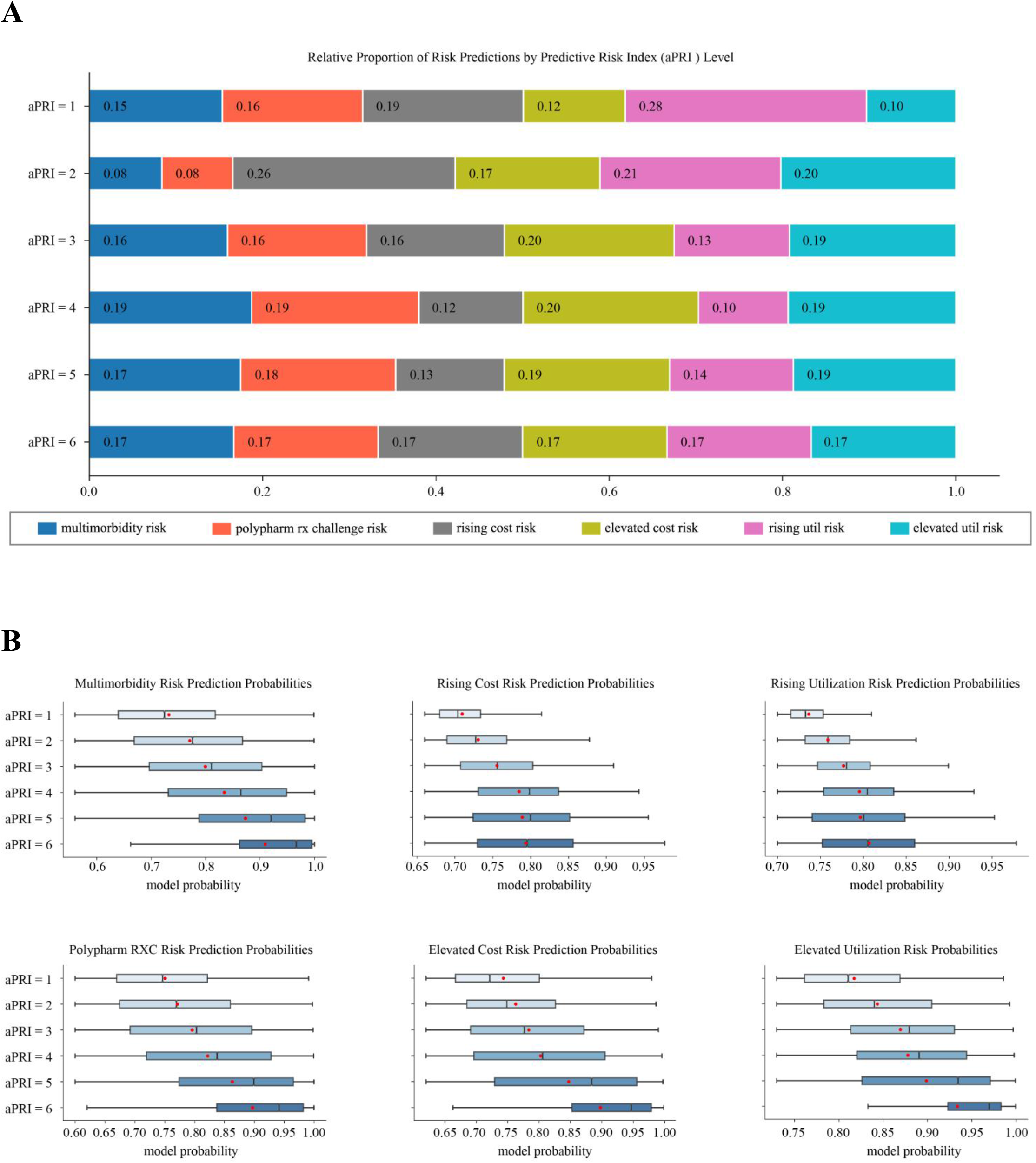
Prevalence of each input model’s “at-risk” (class 1) predictions within aPRI levels 1-6 and distribution of input model’s class 1 probabilities by aPRI level. All characterization was done on holdout data members after calculation of their aPRI values. **A**. Relative prevalence of each input model prediction within aPRI levels 1 – 6. **B**. Notched box plot distributions of input model probabilities for each aPRI level (from 1 – 6). Notch displays 95% confidence interval around the median, red dot indicates average (mean), and outliers are not shown.

For a simplified approach for population level stratification, we can group members into low risk, rising risk, and high risk based on their aPRI levels (see **Figure 6A**). Using this approach, > 80% of holdout dataset members fall into a low risk (aPRI = 0) or rising risk (aPRI = 1-2) category. Less than 20% of members may be considered “high risk” and have aPRI levels of 3 or higher. Across each of these risk groups, age, cost, utilization, and chronic disease count increase on average in support of their “risk labels”. Specific targeting strategies/risk management actions may be employed for each risk group as indicated in **Figure 6A**, with the goal being a reduction in aPRI level year-over-year (especially for high risk members). Depending on population characteristics, organizational resources for member outreach, and real world evidence (i.e., results of outreach efforts at each aPRI level), one may opt to refine further the aPRI cut-points used to define each risk group.

**Figure 6:**
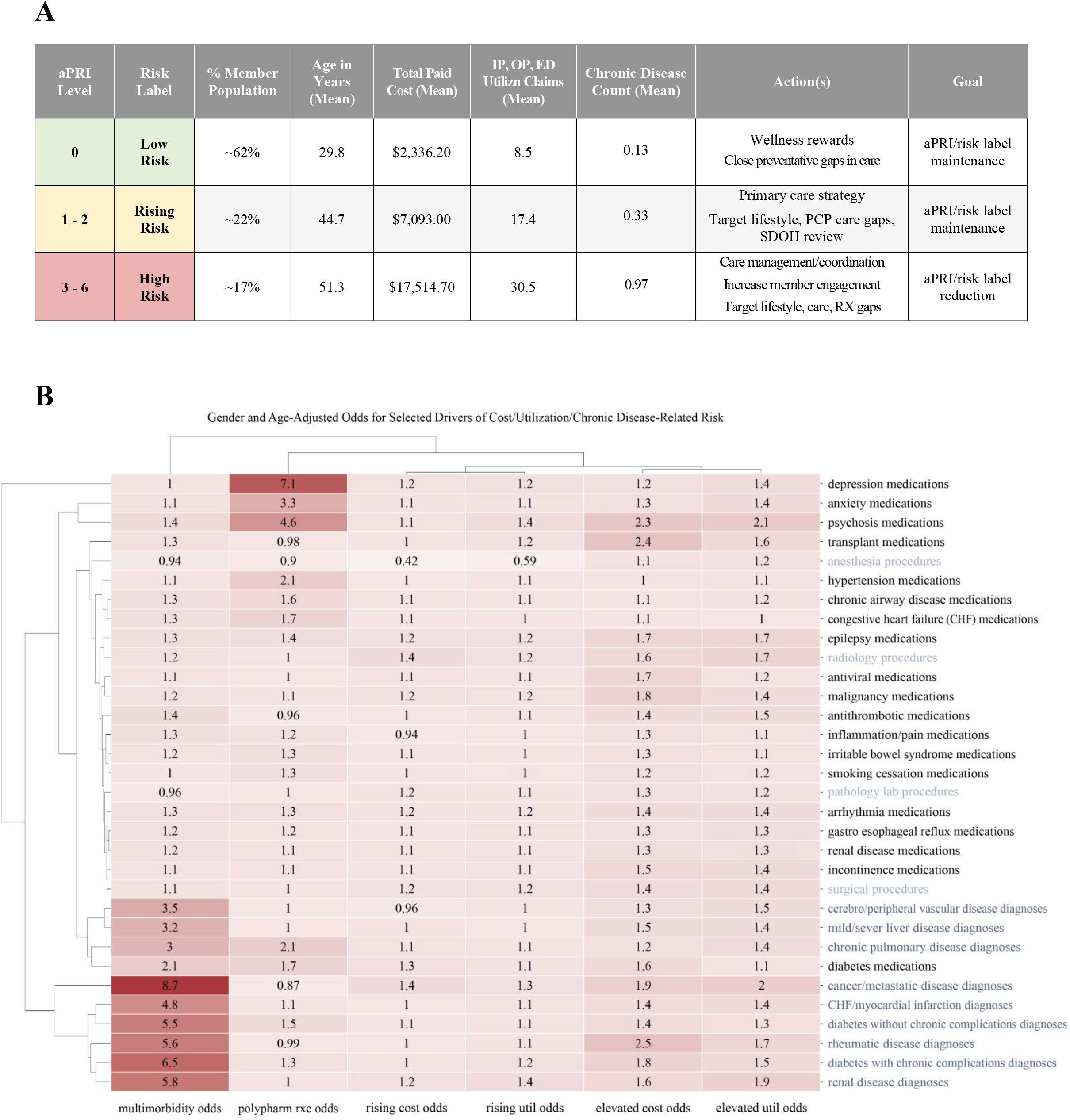
Risk groups based on aPRI level and drivers of each aPRI input modeling “risk outcome”. **A**. Risk groups based on aPRI level in holdout data – relative prevalence and group characteristics (average values) for age, costs, utilizations, and Charlson Comorbidity Index-based comorbidity counts. **B**. Age and gender adjusted odds ratios for selected drivers of modeling outcomes visualized via clustered heat map analysis.

We also identified selected drivers of each of the six risk outcomes for designing additional targeted actions by a member. For this, we extracted age and gender-adjusted odds ratios associated with medications, diagnoses, and procedures against the six outcomes used for predictive modeling; we limited ourselves to these features as we deemed these as more actionable compared to others (e.g., cost and/or utilization-based features). All features associated with at least a 25% increase or decrease in odds were visualized via clustered heat map analysis (**Figure 6B)**. Interestingly, there are both overlapping and unique drivers for each of the risk outcomes, again supporting the complementarity of the input predictions/modeling outcomes. For example, several drivers were identified for multimorbidity risk, at least some of which are also associated with increased odds of polypharm rx challenge (e.g., chronic pulmonary disease diagnosis, diabetes medications intake, and diabetes without chronic complications diagnoses) or elevated cost/utilization risk (e.g., rheumatic disease, renal disease diagnoses). By contrast, intake of medications related to specific behavioral health conditions (anxiety, depression) was primarily associated with increased odds of polypharm rx challenge. Taken together, these drivers can also be leveraged as part of member-level risk stratification/management strategies; actions can include providing targeted support for those on medications for behavioral health and/or prioritizing addressing gaps in care/treatments for other drivers most strongly associated with risk outcomes.

## DISCUSSION

As key components of population health management (PHM) strategies, various predictive/non-predictive approaches are used to identify members at the highest risk/need for targeted management [1-4]. In spite of the multiple approaches currently used for PHM, limitations exist in the identification of members at risk. At least some of these limitations stem from a primary focus on identifying “high cost high need” (HCHN) members, the limited risk measures (primarily cost-based) used to identify HCHN members, and the relatively sparse attention to identifying those with lower risk but who might be potentially more impactable (e.g., those with rising cost, rising utilization, and/or complex treatment patterns). Additional limitations arise from the challenge associated with integrating the multiple approaches typically used in health systems.

To address the above limitations and enhance the breadth of coverage for members at risk, we developed an accretive strategy based on integrating six complementary predictive models built using nationwide administrative claims. The six models not only capture HCHN members across disparate risk measures (elevated cost risk, elevated utilization risk, and multimorbidity risk) but also identify those who may be potentially more impactable by predicting members with rising risk/complex treatment patterns (rising cost risk, rising utilization risk, polypharmacy rx challenge risk). To support integration across model predictions and identify their key drivers for targeted management strategies, we used a common model training cohort of 8.97 million members (continually enrolled over 2 years in medical and pharmacy coverage) and a common set of 101 predictors (features). Our strategy integrates across model predictions and yields a composite risk score for each member – referred to as the <u>a</u>ccretive <u>p</u>redictive risk index (aPRI) – that is an unweighted sum of each of the six predicted risks. Based on results against a holdout dataset (of 2.99 million members from nationwide administrative claims), aPRI not only captures a broader set of members at risk but also enables their stratification across multiple risk measures. Greater than 80% of members grouped into a low risk (aPRI = 0) or rising risk category (aPRI = 1 – 2) and ∼17% grouped into a high-risk category (aPRI = 3-6). These risk categorizations are supported by increasing age, costs, utilization, and chronic disease burden across each group, underlying aPRI levels, and increasing prediction probabilities (especially for HCHN predictions) at higher aPRI levels.

At least a few limitations of our work deserve further mention. First, we selected 6 disparate predictive models for cost, utilization, and chronic-disease related burden, to the increased breadth of predictions for members at risk. However, our models have limited recall/sensitivity for rising cost risk and rising utilization risk predictions. Additional features may be needed for improving these models – but they were either not represented in our CCAE database (e.g., social determinants of health) or removed during data de-identification (e.g., racial information, zip code details, etc.). Second, almost certainly, the six predictive models do not capture all the sources of risk that are relevant for population health management. For example, identifying members with avoidable and/or excess inpatient/emergency department use is commonly done to identify utilization-based risk [1 6], but we did not include predictions for specifically targeting these members. On the contrary, our utilization risk predictions are skewed towards primarily outpatient users (since they account for the vast majority of utilization claims and may be more representative of population health). Other unforeseen gaps may also arise from our definitions of risk used. In theory, more predictive models can be added to our aPRI framework to address these issues – as long as they increase the identification of members at risk and support their stratification. Alternatively, higher risk members identified based on their aPRI levels can be further processed for additional predictive/non-predictive insights and leverage these additional insights for developing targeted management strategies. As a way to support the latter strategy, we have developed multiple approaches in-house to identify members with the potential for various patterns of inpatient and/or emergency department use, specialty drug use, complications for various chronic diseases, progress to new/more severe chronic diseases, polypharmacy/Rx challenge risk by condition etc.

In spite of the above limitations, our accretive predictive modeling framework/strategy not only captures a broader set of members at risk than current approaches but also enables their stratification across multiple risk measures. By assigning a member-level score, our approach also allows one to monitor year-over-year changes in member risk status and identify/prioritize actions or predictive model outputs that are most amenable to risk reduction. Finally, because members with multiple risk scores can exist within the highest risk level (e.g., high risk members as defined in this study have aPRI levels 3-6), one can prioritize outreach efforts based on a combination of factors that include member’s aPRI level, organizational capacity for risk management, and propensity to succeed based on member-level input risk predictions.

## CONCLUSION

We have developed an accretive predictive modeling framework for population-level risk stratification and management.

## Data Availability

The data used for this manuscript are commercially licensed and not publicly available.

## ACKNOWLEDGEMENTS

We appreciate the graphic design inputs from Elizabeth Fultz, Kaylynn Horrigan and their assistance with figure formatting.

## DATA AVAILABILITY

The data used for this manuscript (CCAE) is commercially licensed and not publicly available.

